# Treatment Noncompliance Level Among Patients with Type 2 Diabetes Mellitus: A Hospital Based Cross-Sectional Study in Bangladesh

**DOI:** 10.1101/2022.01.12.22269163

**Authors:** Ishtiakul Islam Khan, Orindom Shing Pulock, Biddhuth Barua, Taslima Ahmed Dola, Pratik Chowdhury, Tapan Seal, Sanzida Salekin, Md. Tarek Ul Quader, Ambarish Mitra, M A Hassan Chowdhury, Sayeda Rumman Aktar Siddiqui

## Abstract

**Introduction:** The consequence of good diabetic treatment depends on the patient’s commitment to a large degree. Noncompliance leads to inadequacy of metabolic control, which strengthens the advancement and speeds up diabetic complications.

**Methodology:** This descriptive cross-sectional study was conducted at Medical Center Hospital, Chattogram, Bangladesh. The study included two hundred and fifty-nine patients with T2DM. Data regarding sociodemographic factors, patient’s characteristics, medication factors, physician-related factors, and noncompliance were collected using a pretested and structured questionnaire. Treatment adherence was assessed by Morisky Medication Adherence Scales (MMAS-8). Data analyses were conducted on SPSS v23.0 Software. The study’s main goal was to assess the treatment noncompliance level among patients with type-2 diabetes mellitus (T2DM) in Bangladesh.

**Result:** The majority of the participants (56%) were in the 40-45 years of age group, followed by 32% in the older age group (≥60 years), and 62.5% of them were male. One hundred and sixty-eight (64.86%) patients were considered low adherent as per the response of the MMAS-8 scale (score <6), followed by 57 (22.0%) patients were regarded as high adherent (score 8) and 34 (13.13%) patients were considered medium adherent (score 6-7) to treatment. Observing the frequency distribution for noncompliance, financial concerns (32.3 %), forgetfulness (27.7%), a busy daily schedule (17.7%), and fear of antihyperglycemic drug side effects were all identified as significant explanations. On multivariate analysis, participants aged 60 years or more, monthly family incomes of <30,000BDT or 30,000 – 50,000 BDT, smoking, and uncontrolled glycemic status showed higher chances of noncompliance than their counterparts.

**Conclusion:** Patient counseling and awareness programs may enhance treatment adherence among people with T2DM. Our findings will help physicians and public health workers to develop targeted strategies to increase awareness of the same among their patients.

**Highlights:**

1. One of the most preventable causes of treatment failure is noncompliance with treatment.
2. Age group, educational level, and monthly family income had a significant association with the compliance to treatment in diabetic patients
3. Majority of the respondents (64.86%) were low adherent to the medications as per MMA-8 scale.
4. Financial issues (32.3 percent), forgetfulness (27.7%), a hectic daily schedule (17.7%), and fear of antidiabetic medicine side effects were all cited as key causes for noncompliance with antidiabetic treatment.
5. Aged participants (≥ 60 years), monthly family income of (30,000-50,000) BDT, uncontrolled glycemic status and smokers portrayed higher frequency of noncompliance.

## Introduction

Compliance is typically known as the degree to which a patient’s behaviour and action correlate with the healthcare provider’s health and medical consultation and recommendations (taking medication, accomplishing behavioural modifications, receiving preventive tests, or sustaining consultation with physicians) [1]. Non-compliant patients are those whose pattern of seeking treatment or maintaining is inconsistent with a health care provider [2]. Patient noncompliance is a crucial health management matter which eventually suggests a preeminent challenge to prosperous healthcare service. It is believed as the most common cause of treatment failure and requires constant motivation from the patients. The result of successful diabetic care outcomes depends on the patient’s adherence at a significant level. Noncompliance leads to a lack of metabolic control, which contributes to the development and acceleration of diabetic complications [3].

The pervasiveness of type 2 diabetes mellitus (T2DM) is soaring worldwide and has become a dominant public health burden. In 2017, approximately 462 million individuals were affected by type 2 diabetes, corresponding to 6.28% of the world’s population. The global prevalence is projected to increase to 7079 individuals per 100,000 by 2030, reflecting a continued rise across all regions of the world [4]. Most forms of diabetes mellitus are of type 2, and the most significant number of people with this condition are between 40 and 59 years of age [5]. The increase in type 2 diabetes is linked to obesity, high blood pressure, and a growing elderly population. Nonetheless, despite solid clinical guidelines for diabetes, the routine of following a healthier lifestyle, adherence to balanced diets, and exercise is minimal. That is why diabetic patients need much more care and follow-up on the diagnosis and recommendations of a physician to ensure their appropriate compliance [2-4].

Many research investigated medication adherence for different conditions, and adherence was consistently found to be suboptimal. Many factors may affect patient adherence to drug therapy. Carelessness, absentmindedness, and casual activities due to a lack of self-discipline, insufficient intelligence, or a courageous philosophy regarding the consequences of diabetes are frequently projected justifications related to nonadherence with oral medication regimens. Throughout developing countries, the rate of noncompliance with long-term care among patients with chronic diseases is around 50% [6], but a higher rate might be observed among developed countries.

Diabetes is a significant public health issue with a high economic burden in Bangladesh. In a sample of 56L452 individuals, the pooled prevalence of diabetes in the general population was 7.8% (95% CI: 6.4–9.3) in Bangladesh, with a significant difference between rural and urban areas. The main factors of diabetes include urbanization, increasing age, and hypertension [7]. The development and improvement of interventions toward better control of T2DM and the prevention of its complications are vital requirements for the country. Without these, soon, the private and public financing of diabetes treatment will be severely constrained, representing a health threat for the Bangladeshi population [8].

Literature search divulged numerous pieces of evidence of variability in adherence to treatment care by age, ethnicity, comorbidity status, socioeconomic status, but most studies conducted had investigated compliance within a given disease state [9–11]. Few have tested adherence across multiple conditions to decide if correlations are compatible between adherence and patient characteristics. But a glaring shortage still exists in the literature about the current treatment compliance level among the diabetic people in the Bangladeshi population. No reliable evidence on the pattern of treatment compliance on antidiabetic drug/treatment among the T2DM in the country is available. The current study focused on and prioritized the current treatment noncompliance level among T2DM patients. It is anticipated that the essential findings of this proposed study would figure out patients’ knowledge about the benefit of following treatment and their attitude towards the currently prescribed treatment. The proposed research tried to overview the current status of diabetic patients’ treatment adherence level and associated factors influencing stoppage or discontinuing the treatment protocol.

Becoming one of the four main types of non-communicable diseases (NCDs), diabetes contributes the most to morbidity and mortality in the world. It may have significant implications for health that impact life expectancy. Life expectancy and morbidity of patients mostly depend on their quality of lifestyle, maintenance as well as following advised treatment. Poor health quality of life begins when glucose levels of a diabetic patient are not managed effectively when they continue smoking, avoid physical exercise, and do not follow treatment properly. Continuation of this lifestyle pattern will ultimately increase the risk of worsening the condition and shorten the life expectancy by developing stroke, heart attack, chronic kidney disease, neuropathy, visual impairment, and amputations. A study in Canada calculated that the disease caused an average reduction of 6 years in females and 5 years in males [12]. A current study of T2DM patients visiting an urban clinic in the capital Dhaka found that nearly two-thirds of patients had uncontrolled diabetes [13]. Additionally, another different study reported poor lifestyle and medication adherence among patients with T2DM in Bangladesh resulting in poor overall quality of life [14].

Mann et al. (2009) conducted a cross-sectional analysis to identify the potentially modifiable patient condition and drug beliefs associated with poor adherence to medication among people with diabetes which portrayed claiming to have diabetes only when becomes elevated, medication avoidance habit, adverse effects related worries, lack of self-confidence and feeling medicines are hard to take [15] were discovered associated with poor medication adherence. But some countries like Ethiopia depicted high compliance (85.1%) among diabetes patient and education, duration of diabetes and knowledge about DM and its medications are significant associated factors related to adherence. [16] A study by Mumu and Saleh portrayed that most of the T2DM patients remained non-adhere to their diet (88%), foot care (70%), routine blood glucose testing (32%) and exercise (25%). [17] Another study by Saha et. al [18] found out that in Bangladesh 44% of the patients were considered moderately adherent and 19% were poorly adherent while most of their (75%) respondents were female. Moreover, significant (p< 0.05) relation between Noncompliance and quality of life in diabetic patients in Bangladesh. [19]

Adherence to medicines and lifestyle changes has an important impact on the outcome of diabetes treatment along with their quality of life. Furthermore, non-adherence to diabetes medication remains an unresolved issue, which can lead to several expensive and life-threatening complications and the experience of the patient is often ignored in the scheme of the treatment. It is therefore important to understand the perspective of patients on diabetes, its medicines, and the significance of adherence to glycemic control medications to facilitate effective and optimal diabetes treatment. Bangladesh lacks reliable research and data on the viewpoint of patient’s compliance to treatment and additional advice. Therefore, the present study was designed to address some of these knowledge gaps and to better understand the current compliance of T2DM patients in urban Bangladesh.

## Methodology

### Study design

This is a descriptive type of cross-sectional study took place in a tertiary care medical center that provide a comprehensive healthcare service for the people of Chattogram, the southern part of Bangladesh. The main aim of this study is to describe the current treatment noncompliance level among Type-2 diabetes mellitus patients and its associated factors. The study population comprised the diagnosed patients with T2DM attending Indoor patient department (IPD) and outdoor patient department (OPD). The study was conducted from April 2020 to August 2020. Data were collected for three months within the five months study period. Convenient type of sampling method was used and data was collected by face-to-face interview with maintaining proper safety measures for limiting COVID-19 exposure. Since prevalence of type-2 diabetes is low among < 30 years aged group, patient aged 30 and above and patients who had been diagnosed with diabetes for at least six months from the date of interview were included in the study. Pregnant or lactating women up to 12 weeks post-partum were excluded due to the possible pregnancy-related impaired glucose tolerance status in this group.

### Sample size

In total, two hundred and sixty (260) diabetic patients from the selected hospital were included by assuming a confidence interval of 95% and a power of 80%, prevalence of 21%. According to a study conducted by Shaha et al., the prevalence of treatment noncompliance among T2DM patients in the Bangladeshi population was found at 21% [18]. For calculation of the sample size, the following formula was used:

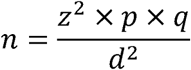

Here,

n = desired sample size

z = standard normal deviate; set at 1.96, which correspond to 95% confidence level. P = 0.21 (prevalence of treatment Noncompliance among T2DM patients among Bangladeshi population is 21%) [18]

q = 1-p =1-0.21 = 0.79

d = Allowable margin error = 5% =0.05

So,

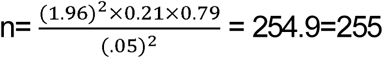

The final sample size was 260 patients.

### Data Collection Tools

Data for this descriptive cross-sectional study were collected via a structured pretested questionnaire. It had three sections: The first section included questions regarding sociodemographic factors, the second section contained questions regarding clinical factors, and the third section was the adherence assessment tool known as Morisky Medication Adherence Scales (MMA-8) [20]. The MMAS-8 has far greater sensitivity and specificities of 93% and 53% and the alpha value of 0.83 of Cronbach respectively [21]. The first seven items are dichotomous categories with either yes or no, and the last item examines how often the respondents skip the daily doses of medicines with five different responses and their corresponding scores. The MMAS-8 is a self-report questionnaire with 8 questions (items) whose wording of the questions/items are formulated to avoid answering “yes” to questions regardless of their content. Items 1 through 7 have response choices “yes” or “no” whereas item 8 has a 5-point Likert response choice. Each ‘‘no” response is rated as ‘‘1” and each ‘‘yes” is rated as ‘‘0” except for item 5, in which each response ‘‘yes” is rated as ‘‘1” and each ‘‘no” is rated as ‘‘0”. For item 8, if a patient chooses response ‘‘0”, the score is ‘‘1” and if they choose response ‘‘4”, the score is ‘‘0”. Responses ‘‘1, 2, 3” are respectively rated as ‘‘0.25, 0.75, 0.75”. Total MMAS-8 scores can range from 0 to 8 and have been categorized into three levels of adherence: high adherence (score = 8), medium adherence (score of 6 to < 8), and low adherence (score< 6).

Random Blood Sugar Measuring Machine was used in the survey day, to measure the random blood sugar level (RBS) of the diabetic patients. Measuring tape and weight machines were used to measure height and weight to detect the current Body mass index (BMI). The study was conveyed after obtaining permission from the concerned authorities and the participants as well.

### Data Management & Analysis Plan

After the collection of data, they were checked and verified for consistency and reduction of errors. Data entry and analysis were completed by Statistical Packages for Social Sciences (SPSS) version 23.00. The continuous variables were categorized and descriptive statistics were calculated (presented as frequencies). Various variables such as demographics (age categorized as <40 years, 40-59 years or ≥60years and gender), monthly income categorized as <30,000 Bangladeshi Taka (BDT), 30,000-50000 BDT, and >50,000 BDT, and education split into two categories; less than higher secondary education and education up to higher secondary and above), smoking status (never smoker and smoker), marital status (married and unmarried which included separated, divorced, widowed and widower) comorbidities, were the independent variables. Univariate analyses (Chi-square or Fisher’s exact test) were performed to assess the association of each of the independent variables and treatment compliance (compliant versus non-compliant). To determine which factors were predictive of non-compliant when adjusted for other predictors, a multiple binary logistic regression was performed. Explanatory variables were selected using liberal criteria (P <0.05) for inclusion in the multivariate regression model. Odds ratios (ORs) and 95% confidence intervals (CIs) were calculated for all comparisons. For all statistical analyses, significance was accepted as p <0.05. A p-value of <0.05 was regarded as statistically significant for all analyses.

## Result

This study included 260 patients, one of whom did not have complete information regarding medication adherence. Therefore, finally, 259 patients were analyzed. The sociodemographic characteristics of the participants are summarized in Table 1. The majority of the participants were 40-45 years of age, followed by 32% in the older age group (≥60 years). More than 62% (62.5%) of them were male. Most participants were married (94.2%) and educated up to the higher secondary school level and above (67.2%). The majority of the patients’ (55.2%) monthly income was below BDT 30,000 **(Table 1)**.

**Table 1:**
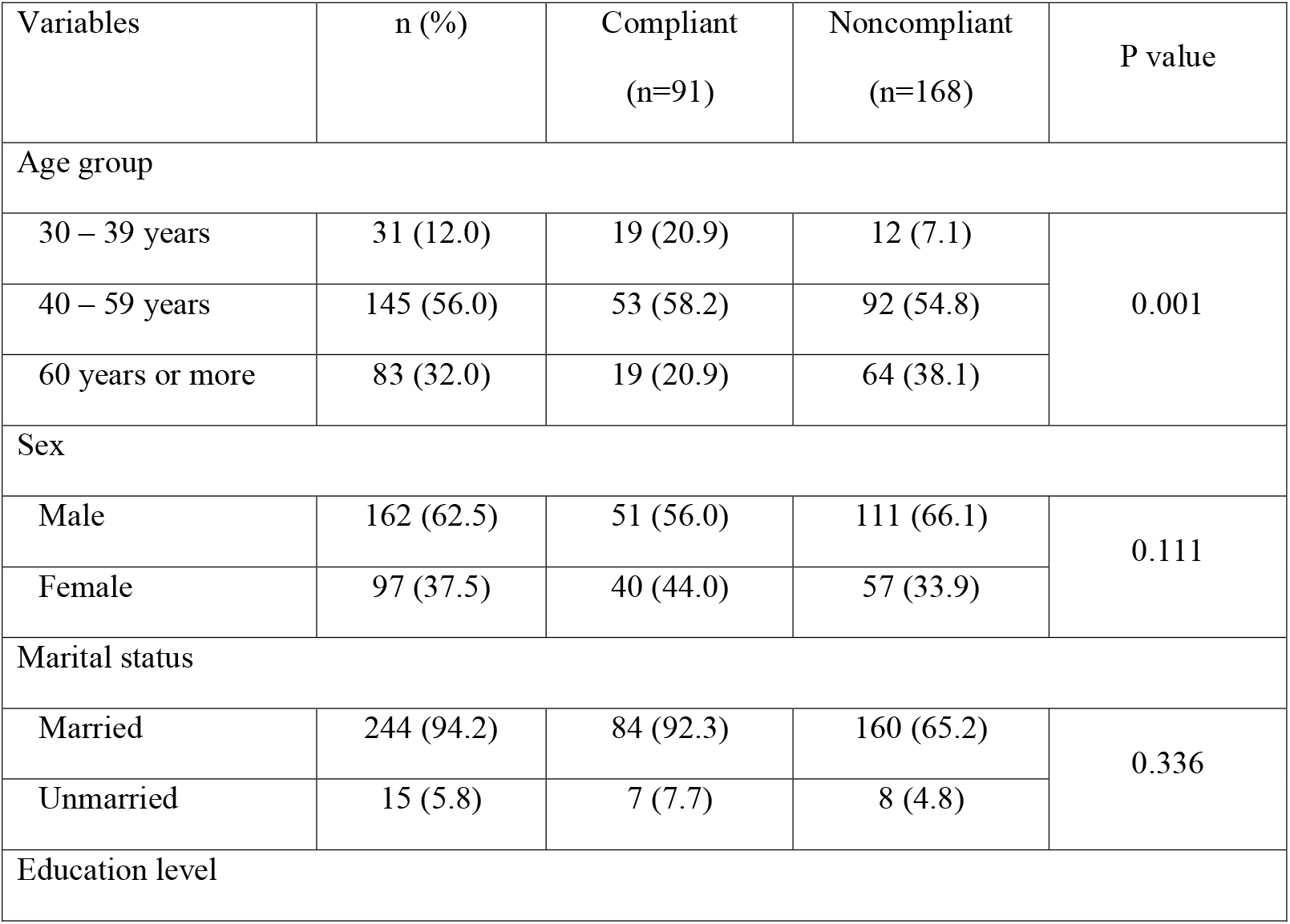
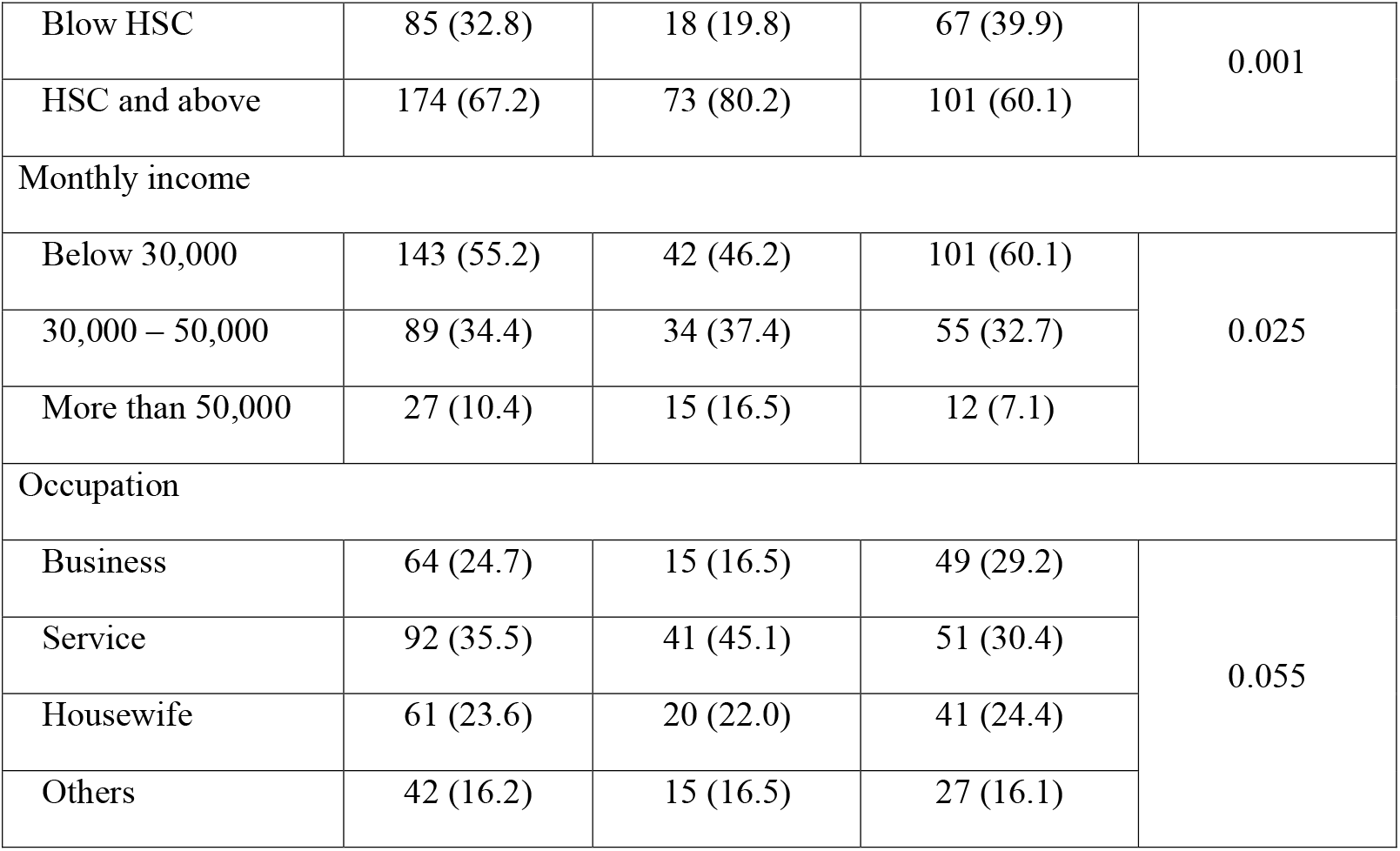
Socio-demographic characteristics of the study participants (n=259)

| Variables | n (%) | Compliant<br>(n=91) | Noncompliant<br>(n=168) | P value |
| --- | --- | --- | --- | --- |
| Age group |  |  |  |  |
| 30 – 39 years | 31 (12.0) | 19 (20.9) | 12 (7.1) | 0.001 |
| 40 – 59 years | 145 (56.0) | 53 (58.2) | 92 (54.8) |  |
| 60 years or more | 83 (32.0) | 19 (20.9) | 64 (38.1) |  |
| Sex |  |  |  |  |
| Male | 162 (62.5) | 51 (56.0) | 111 (66.1) | 0.111 |
| Female | 97 (37.5) | 40 (44.0) | 57 (33.9) |  |
| Marital status |  |  |  |  |
| Married | 244 (94.2) | 84 (92.3) | 160 (65.2) | 0.336 |
| Unmarried | 15 (5.8) | 7 (7.7) | 8 (4.8) |  |
| Education level |  |  |  |  |
| Blow HSC | 85 (32.8) | 18 (19.8) | 67 (39.9) | 0.001 |
| HSC and above | 174 (67.2) | 73 (80.2) | 101 (60.1) |  |
| Monthly income |  |  |  |  |
| Below 30,000 | 143 (55.2) | 42 (46.2) | 101 (60.1) | 0.025 |
| 30,000 – 50,000 | 89 (34.4) | 34 (37.4) | 55 (32.7) |  |
| More than 50,000 | 27 (10.4) | 15 (16.5) | 12 (7.1) |  |
| Occupation |  |  |  |  |
| Business | 64 (24.7) | 15 (16.5) | 49 (29.2) | 0.055 |
| Service | 92 (35.5) | 41 (45.1) | 51 (30.4) |  |
| Housewife | 61 (23.6) | 20 (22.0) | 41 (24.4) |  |
| Others | 42 (16.2) | 15 (16.5) | 27 (16.1) |  |

Only 23.9% of patients had no other comorbid conditions, but 39% had more than one comorbid condition. Near about three-fourths of the participants (74.1%) reported having a positive family history of T2DM. Nearly one-fourth (23.9%) were smokers, and more than three-fourths of the participants were overweight **(Table 2)**.

**Table 2:** Clinical characteristics of the study participants (n=259)

| Variables | n (%) | Compliant<br>(n=91) | Noncompliant<br>(n=168) | P value |
| --- | --- | --- | --- | --- |
| Comorbidity |  |  |  |  |
| No | 62 (23.9) | 32 (35.2) | 30 (17.9) | <0.001 |
| One | 96 (37.1) | 38 (41.8) | 58 (34.5) |  |
| More than one | 101 (39.0) | 21 (23.1) | 80 (47.6) |  |
| Family history of DM |  |  |  |  |
| Absent | 67 (25.9) | 19 (20.9) | 48 (28.6) | 0.177 |
| Present | 192 (74.1) | 72 (79.1) | 120 (71.4) |  |
| Smoking status |  |  |  |  |
| Non-smoker | 197 (76.1) | 76 (83.5) | 121 (72.0) | 0.039 |
| Smoker | 62 (23.9) | 15 (16.5) | 47 (28.0) |  |
| BMI category |  |  |  |  |
| <25.0 kg/m<sup>2</sup> | 61 (23.6) | 22 (24.2) | 39 (23.2) | 0.862 |
| ≥25.0 kg/m<sup>2</sup> | 198 (76.4) | 69 (75.8) | 129 (76.8) |  |

Disease duration was less than ten years in the majority of the patients (82.2%), and 69.2% were on oral hypoglycemic agents only. One-third (33.2%) of the patients had uncontrolled glycemic status at data collection **(Table 3)**.

**Table 3:** Distribution of the patients according to present treatment-related factors (n=259)

| Variables | n (%) | Compliant<br>(n=91) | Noncompliant<br>(n=168) | P value |
| --- | --- | --- | --- | --- |
| Duration of diabetes |  |  |  |  |
| < 10 years | 213 (82.2) | 79 (86.8) | 134 (79.8) | 0.156 |
| 10 years or more | 46 (17.8) | 12 (13.2) | 34 (20.0) |  |
| Current treatment |  |  |  |  |
| Diet and exercise only | 5 (1.9) | 4 (4.4) | 1 (0.6) | 0.053 |
| OHA only | 179 (69.2) | 70 (76.9) | 109 (64.9) | 0.045 |
| Insulin only | 26 (10.0) | 8 (8.6) | 18 (10.7) | 0.623 |
| Combinations of OHA and insulin | 49 (18.9) | 9 (9.9) | 40 (23.6) | 0.006 |
| RBS, mmol/l |  |  |  |  |
| Controlled (<11.1) | 173 (66.8) | 73 (80.2) | 100 (59.5) | 0.001 |
| Uncontrolled ($\geq 11.1$) | 86 (33.2) | 18 (19.8) | 68 (40.5) | |

Out of 259 patients, 168 (64.86%) were considered low adherent as per the response of the MMAS-8 scale (score <6). Only 57 (22.0%) patients were considered high adherent (score 8) and 34 (13.13%) patients were considered medium adherent (score 6-7) **(Fig 1)**. For further analysis, participants of this study were grouped as treatment non-compliant (low medication adherence) and compliant (medium/high medication adherence). Fig 2 reveals that 168 (64.9%) participants were classified as non-compliant, while the remaining 91 (35.1%) were classified as complying.

**Fig 1:**
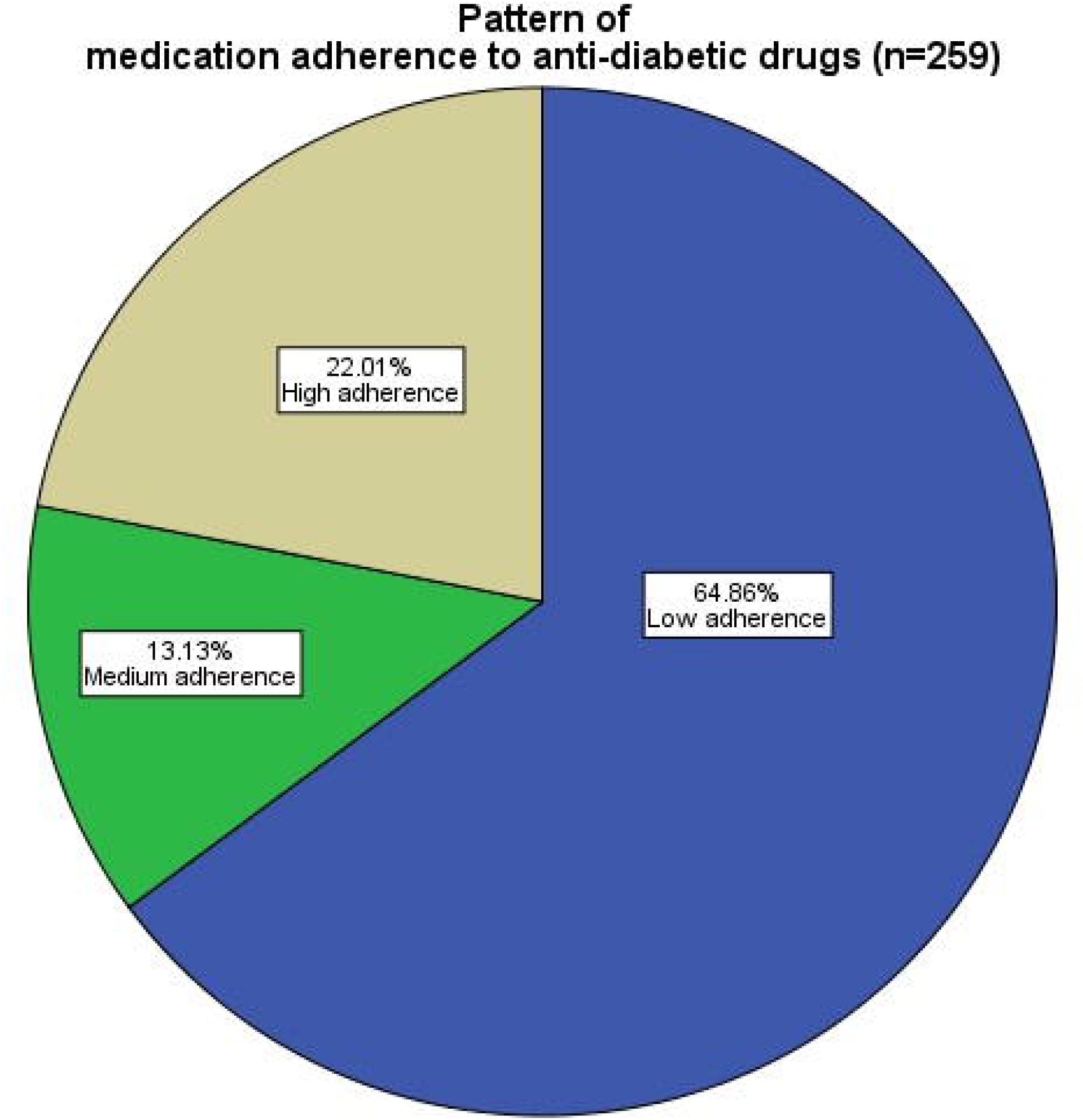
Distribution of patients according to the pattern of medication adherence to anti-diabetic drugs.

**Fig 2:**
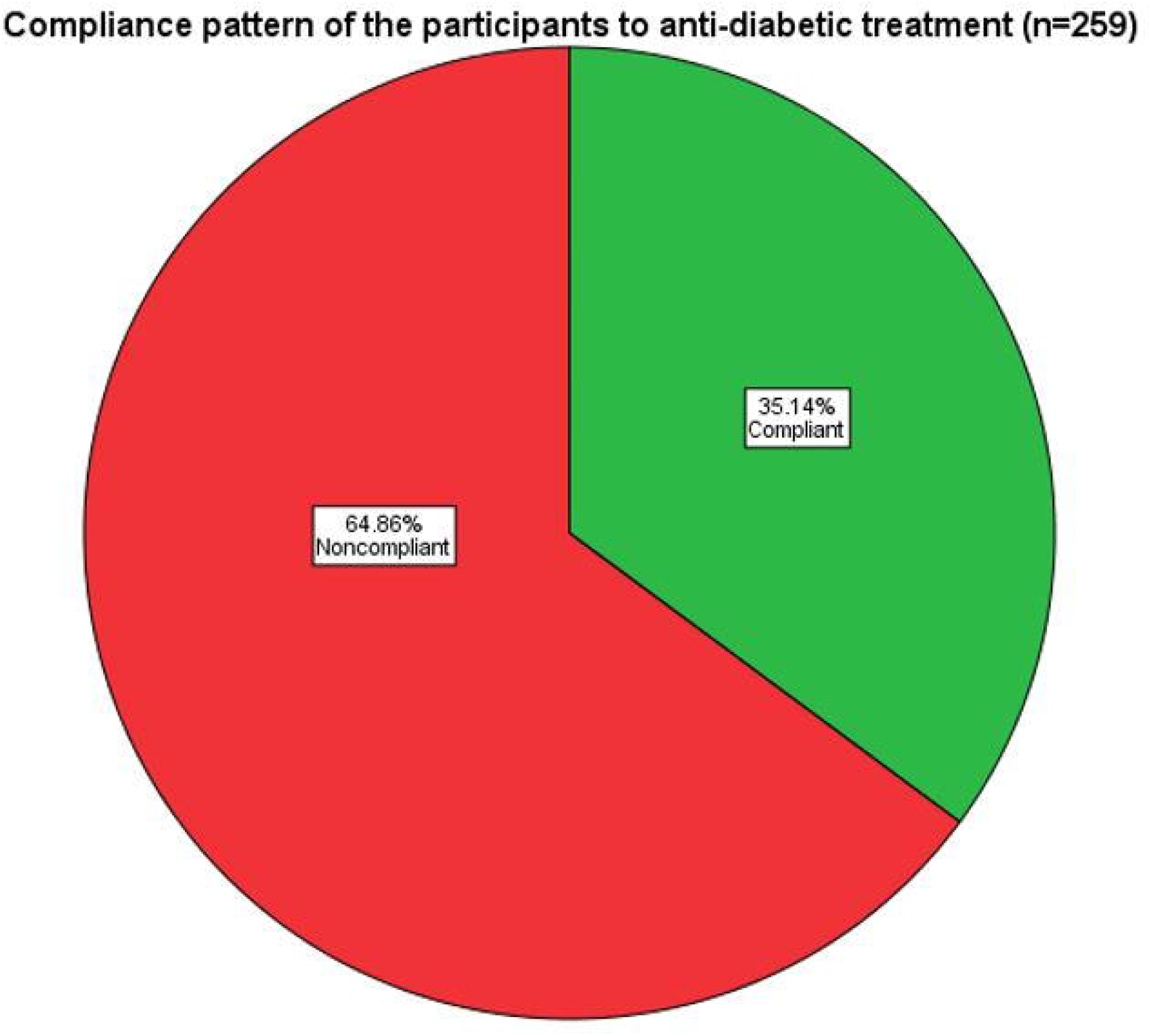
Distribution of patients according to pattern of compliance to anti-diabetic treatment.

Financial problems (32.3%), forgetfulness (27.7%), busy daily schedule (17.7%), and fear of side effects of antidiabetic drugs were considered as major reasons for being non-compliant with antidiabetic treatment **(Fig 3)**.

**Fig 3:**
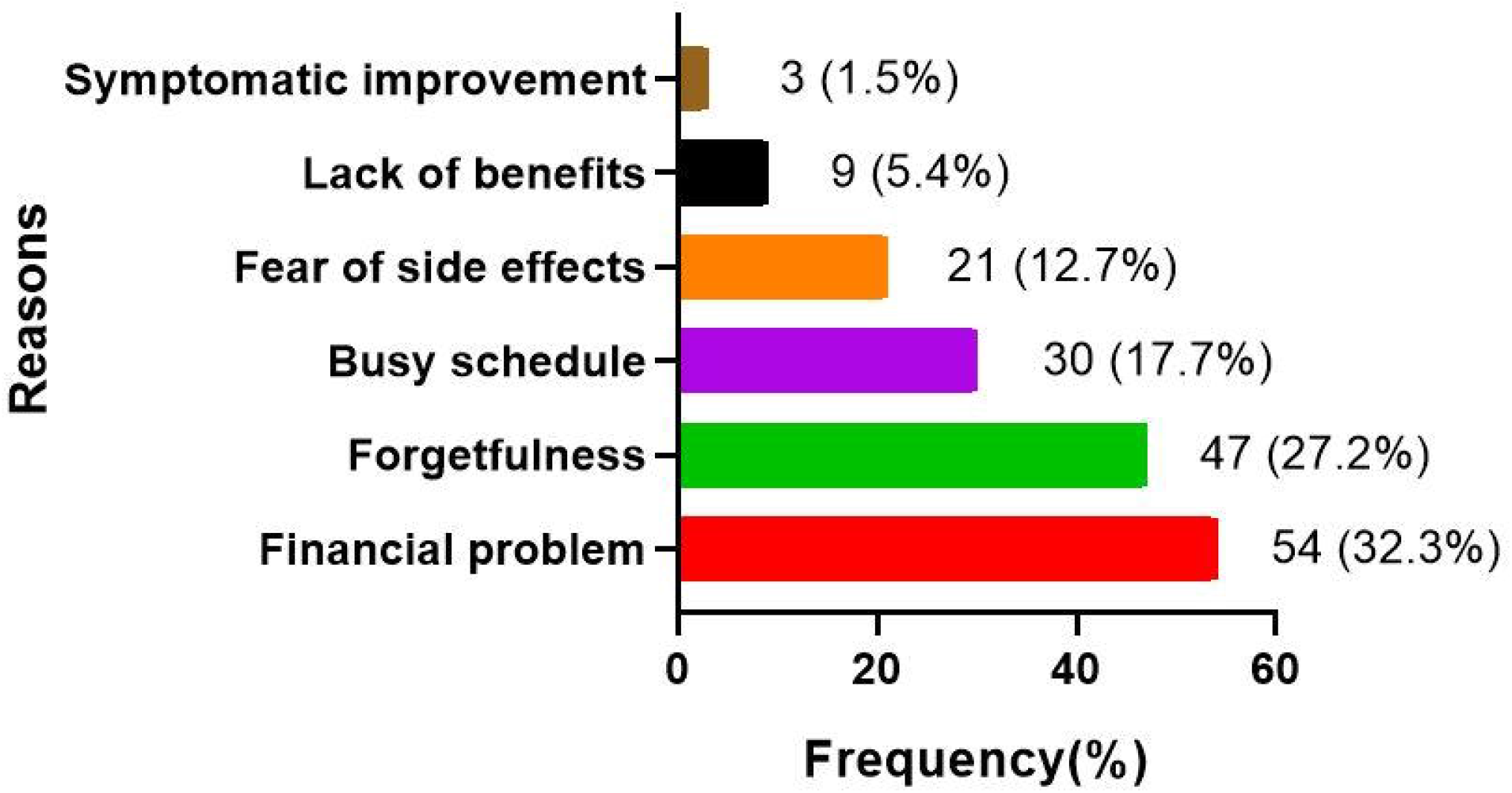
Reasons for not taking anti-diabetic treatment in non-compliant patients (n=168).

Age group, educational level, and monthly family income had a significant association with the compliance to treatment in diabetic patients **(Table 1)**. On the other hand, gender, marital status, and occupation had no association with compliance to treatment in diabetic patients. The frequency of patients with more than one comorbid condition was higher in the non-compliant group (p<0.001) and in smokers (p=0.039) compared to their counterparts (Table 2). On the other hand, though the frequency of non-compliant was higher in patients without any family history of DM and patients with BMI <25 kg/m^2^compared to their counterparts, these differences failed to reach statistical significance (p.0.05).

The study diseases duration category (<10 yrs. versus ≥10 yrs.) had no association with treatment compliance. The current treatment pattern had a significant association with treatment compliance, and noncompliance was associated with the glycemic status of the patients **(Table 3)**.

As per the adjusted bivariate logistic regression analyses, participants aged 60 years or more had 3.83 times (95% CI:1.35-10.85; p=0.012) higher odds of noncompliance than the participants (Table 8). Participants with monthly family incomes of <30,000BDT or 30,000 – 50,000 BDT were 6.08 times (95% CI: 2.19-16.82) and 4.9 times (95% CI: 1.45-11.51) more likely to have noncompliance, respectively, as compared to those with a monthly family income of >50,000BDT. In addition, smokers (OR: 2.1, 95% CI: 1.05-4.59) and had uncontrolled glycemic status (OR: 2.38, 95% CI: 1.10-5.18) showed higher chances of noncompliance compared to their counterparts. **(Table 4)**

**Table 4:** Factors associated with Noncompliance to treatment among diabetes patients.

|  | Unadjusted OR | P value | Adjusted OR (95% CI) | P value |
| --- | --- | --- | --- | --- |
|  | (95% CI) |  |  |  |
| **Age group** |  |  |  |  |
| 30 – 39 years | Reference |  | Reference |  |
| 40 – 59 years | 2.75 (1.24-6.10) | 0.013 | 2.30 (0.96-5.54) | 0.063 |
| 60 years or more | 5.33 (2.20-12.93) | 0.001 | 3.83 (1.35-10.85) | 0.012 |
| **Education level** |  |  |  |  |
| Below HSC | 2.69 (1.48-4.91) | 0.001 | 1.42 (0.71-2.84) | 0.329 |
| HSC and above | Reference |  | Reference |  |
| **Monthly income** |  |  |  |  |
| Below 30,000 | 3.01 (1.29-6.96) | 0.010 | 6.08 (2.19-16.82) | 0.001 |
| 30,000 – 50,000 | 2.02 (0.84-4.83) | 0.113 | 4.09 (1.45-11.51) | 0.008 |
| More than 50,000 | Reference |  | Reference |  |
| **Comorbidity** |  |  |  |  |
| No | Reference |  | Reference |  |
| One | 1.63 (0.86-3.10) | 0.138 | 1.39 (0.68-2.89) | 0.364 |
| More than one | 4.06 (2.03-8.12) | <0.001 | 2.68 (1.21-5.95) | 0.016 |
| **Smoking status** |  |  |  |  |
| Non-smoker | Reference |  | Reference |  |
| Smoker | 1.97 (1.03-3.76) |  | 2.19 (1.05-4.59) | 0.037 |
| **Combination treatment** |  |  |  |  |
| No | Reference |  | Reference |  |
| Yes | 2.85 (1.31-6.18) | 0.008 | 2.03 (0.64-6.47) | 0.230 |
| **Only OHA** |  |  |  |  |
| No | 1.80 (1.01-3.23) | 0.047 | 1.43 (0.78-3.91) | 0.485 |
| Yes | Reference |  | Reference |  |

| **RBS, mmol/l** |  |  |  |  |
| --- | --- | --- | --- | --- |
| Controlled (<11.1) | Reference |  | Reference |  |
| Uncontrolled (≥11.1) | 2.76 (1.51-5.03) | | 2.38 (1.10-5.18) | <0.001 |

## Discussion

T2DM is a chronic disease requiring lifelong treatment, although lifestyle modifications play an important role in diabetes management. This study was conducted to determine treatment noncompliance among T2DM attending a private hospital in Bangladesh. Altogether a total of 259 prescriptions were analyzed during the study period. The present study showed that more than half of the participants with T2DM (64.86%) had low treatment adherence, and another 13.13% had medium treatment adherence. Only 22.0% of patients were revealed as having high treatment adherence in the current study. A previous study in Bangladesh measured medication adherence among patients with T2DM and reported that 20% of the participants were non-adherent to oral medication [19], which is much lower than our findings.

In contrast, Saleh et al. showed higher adherence in their study population, but this might be because they did not measure adherence using a standardized questionnaire [19]. Our findings align with a report from India that used a standard medication-adherence tool and found 51.7% of their participants to have low adherence. Another report from Saudi Arabia reported that only a third of patients had high adherence to their prescribed antidiabetic medications [22, 23]. Moreover, a high prevalence of low-treatment adherence in T2DM patients, as revealed by the current study, is confirmed by another recent study from Bangladesh, which reported that the overall prevalence of low adherence was 46.3% of participants and medium-to-high adherence was 53.7% in patients with T2DM [24].

In the present study, a high proportion (56 %) of patients in this study were in the age group of 40-59 years of age, and the proportion of elderly (age ≥60 years) patients was low (32%). However, older age was a significant predictive factor for treatment noncompliance in the current study. This finding could be because younger patients take better care of their health to ensure a long healthy life, and the elderly seem to fear complications and mortality. This reported adherence rate was consistent with the previous findings [25,26]. Moreover, older patients who suffer from comorbidities should have more medicines on their prescription, and polypharmacy is a potential factor for medication nonadherence [27]. In contrast to these findings, AlQarni et al. reported medication adherence correlated positively with patient age as their findings suggest that patients tended to be more adherent with increasing age [23]. The disparity found can result from the free health scheme, social and psychological support for the elderly [28].

A significant association was observed in the current study between low medication adherence and the education level of the participants. Previous studies have found that patients who have been educated till the primary level showed significantly low adherence to medication [29,30]. The association may be due to the relationship between education and other variables. For example, the educational qualification could determine a patient’s trust in a physician and could further differ according to different levels of education [2].

Low socioeconomic status is a significant factor for poor adherence to medication among diabetic patients [31]. In the current study, participants were categorized into three groups according to their monthly family income. It was revealed that patients with low monthly family income were more likely to be non-compliant to treatment than the participants with high monthly family income. The cost of medication is a militating factor affecting patients’ adherence to their medications. According to a study carried out by Awodele et al., more than half of the patients (51.32 %) viewed their drugs as unaffordable [28]. In the current study, the financial problem was the main reason stated for treatment non-compliant.

It is well known that diabetic patients who smoke are less likely to be active in self-care or comply with diabetes care recommendations [32]. In the present study, diabetic patients who never smoked were 2.19 (95% CI:1.05-4.59) times more likely to have treatment non-compliant than those who never smoked in their lifetime.

The current study found noncompliance to be associated with more than one comorbidity. Patients with more than one additional comorbid condition were 2.68 (1.21-5.95) times more likely to be non-compliant than those with no other comorbid condition. Diabetic patients with multimorbidity had to take multiple medications in addition to antidiabetic medicines. Similarly, Shams et al. also reported that diabetic patients with different associated comorbid conditions like ischemic heart disease, hypertension, and patients taking >4 drugs were more likely to report nonadherence to medication [27].

Importantly, this study demonstrated that treatment compliance plays a vital role in maintaining blood sugar levels within the normal range. This study found patients with uncontrolled glycemic status were 2.38 (95% CI:1.10-5.18) times more likely to be treatment non-compliant than patients with reasonable glycemic control. Similarly, a significant inverse relationship between high adherence scores and lower assayed values of HbA1C and FBS was reported by Rana et al. [25].

Patients with a family history of diabetes were not significantly associated with low medication adherence in this study, similar to a previous study conducted in Pakistan [24]. Reportedly, the family members of diabetes patients are more knowledgeable about diabetes, but they perform more non-supportive behaviors, leading to patients being less adherent to their medication [33].

However, it is to be noted that inconsistencies prevail in the literature regarding the factors associated with treatment compliance [2]. It is attributable to the lack of standard techniques to measure adherence, differences in sample populations, and different definitions of glycemic control.

## Limitations

There are a few limitations to this study. First, this study primarily included subjects residing in urban areas seeking health care at private hospitals within Chittagong city and had access to specialized care and education on diabetes management protocols. Moreover, the sample size was relatively small. Second, it was not possible to collect data on several contributing factors, such as health literacy, food frequency, and pathophysiological factors that could be relevant to medication adherence. Third, patients may have overestimated their medication adherence in their assessments, but these results could not be validated with more accurate adherence measurements. Finally, a cross-sectional study cannot establish the causal factor and does not provide any in-depth information. Regardless of these limitations, this pioneering study in Bangladesh provides a novel country-specific analysis of treatment compliance among patients with T2DM using a standardized assessment tool.

## Conclusion

Despite the public health efforts being made to effectively manage diabetes among the population of Bangladesh, increasing medication adherence is still a key challenge among patients with T2DM in Bangladesh. The factors identified to be associated with low medication adherence among diabetic individuals in the current study included older age, family income of less than 30,000 BDT, education below HSC level, being a smoker, and having more than one comorbid condition. These diabetic patients should be considered at high risk of nonadherence and are likely to require more creative and consistent clinical interventions. These findings will help physicians and public health workers to design innovative interventions to address these and eventually improve medication adherence in Bangladesh.

## Data Availability

All data produced in the present study are available upon reasonable request to the authors

## Abbreviations

BDT: Bangladeshi Taka
BMI: Body Mass Index
IPD: Indoor Patient Department
HSC: Higher School Certificate
MMAS-8: Morisky Medication Adherence Scales
NCDs: Non-Communicable Diseases
OPD: Outdoor Patient Department
RBS: Random Blood Sugar
SPSS: Statistical Packages for Social Sciences
T2DM: Type 2 Diabetes Mellitus

## Acknowledgements

We acknowledge the dedications, commitments and the contributions of the patients who took part in this study. We are grateful for the valuable contribution from the study site management authority for granting us the permission for data collection.

